# Separate functional and structural cerebral mechanisms relate to postanoxic coma recovery

**DOI:** 10.1101/2025.08.13.25333575

**Authors:** Marlous M.L.H. Verhulst, Hanneke M. Keijzer, Ting Mei, Catharina J.M. Klijn, Cornelia Hoedemaekers, Michiel Blans, Bart A.R. Tonino, Frederick J.A. Meijer, Christian F. Beckmann, Rick C. Helmich, Jeannette Hofmeijer

## Abstract

**Background and objectives:** Understanding postanoxic encephalopathy following cardiac arrest is important to provide optimal treatment and predict chances of recovery. Both functional cerebral disturbances (changes in electrophysiological activity or functional connectivity) and structural tissue changes (signs of cytotoxic or vasogenic oedema) have been associated with neurological outcome. We hypothesize that functional and structural changes follow separate pathways towards persistent coma or recovery after a cardiac arrest. We aim to identify common and separate pathways between structural and functional changes using an advanced integration method.

**Methods:** We performed a prospective multicentre cohort study in comatose cardiac arrest patients in three Dutch hospitals. T1-weighted, diffusion-weighted, and resting-state functional MRI scans were collected on day 2-9 after cardiac arrest. Primary outcome measure was neurological outcome at six months defined by the Cerebral Performance Categories (CPC), dichotomized as CPC 1-2 = good, CPC 3-5 = poor. Maps of grey matter volume, Jacobian deformation, mean diffusivity, skeletonized fractional anisotropy, and eight resting-state networks were used as input for a linked independent component analysis. Group differences in subject loadings on the components between individuals with good and poor outcome were calculated. Relationships between functional and structural components were studied descriptively.

**Results:** We included 80 patients (31 with poor outcome). Of twenty identified components, six consisted of functional, seven of structural, and five of both structural and functional information. Four components related independently to outcome: two functional and two structural components. None of the components containing both functional and structural information related to outcome. Patients with good outcome showed higher connectivity in multiple resting-state networks (mainly visual, default-mode, and frontoparietal network) and smaller volumes of and higher diffusivity in the cortex and deep grey nuclei. Signs of functional injury were seen in both outcome groups, while only patients with poor outcome showed signs of structural damage. Forty-three patients showed functional without structural, but no showed structural without functional injury.

**Discussion:** Functional network and structural tissue injury exist independently in postanoxic encephalopathy after cardiac arrest. Our results point towards distinct cerebral mechanisms relating to postanoxic coma outcome, likely including cell swelling, tissue edema, and isolated synaptic failure.

**Clinical trial registration:** Clinicaltrials.gov NCT03308305

## Introduction

Each year, approximately 55 per 100,000 people worldwide have a cardiac arrest, with a 7% survival rate to hospital discharge.^1^ After return of spontaneous circulation (ROSC), the majority of patients initially remains comatose due to postanoxic encephalopathy. Of these patients, admitted to the intensive care unit (ICU), approximately 50% will not survive as a result of irreversible ischemic brain injury.^2^ There are no treatments to improve neurological recovery with unequivocal evidence of efficacy.^3–5^ Postanoxic encephalopathy and subsequent recovery involve a cascade of pathophysiological processes that is important to understand in order to provide optimal intensive care treatment, accurate prediction of recovery, and development of targeted treatment strategies.

Changes in synaptic functioning are the earliest consequence of cerebral ischemia and may occur within seconds, even at moderately reduced perfusion levels.^6,7^ Early synaptic failure mostly results from presynaptic injury with impaired transmitter release.^6^ Disappearance of synaptic activity may be reversible. However, without timely recovery of cerebral perfusion, disturbances of synaptic transmission may become permanent, even with preserved membrane potential.^8^ Several observations under experimental conditions have suggested that ongoing, isolated synaptic failure after an hypoxic insult can result from permanently impaired transmitter release. These observations include decreased postsynaptic potentials when afferent fibres are stimulated with intact responses to exogenous postsynaptic receptor agonists,^6^ intact spontaneous miniature excitatory postsynaptic potentials,^9^ and a preserved function of pyramidal cells to generate action potentials.^10^ Synaptic injury goes along with disturbed EEG patterns,^11^ but will not be visualized with conventional imaging techniques.^6,7^

With continued cardiac arrest, cerebral perfusion levels decrease, leading to the failure of ATP - dependent ion pumps. This disrupts ion gradients across the plasma membrane, altering osmotic pressures and eventually resulting in cytotoxic oedema.^7,12^ As the situation progresses, excitatory neurotransmitter release triggers calcium influx, causing mitochondrial dysfunction and other cellular damage.^12^ Meanwhile, prolonged hypoxia compromises the blood-brain barrier that, together with osmotically active substances accumulating in brain tissue, leads to vasogenic oedema.^13^ Upon restoration of blood flow, additional injury mechanisms such as oxidative stress, inflammation, microvascular damage, and impaired vasoregulation may exacerbate neuronal injury.^12,14,15^ Altogether, these pathophysiological mechanisms may lead to a cascade from cytotoxic oedema to vasogenic oedema to structural brain tissue injury that is classically visualized by various MRI techniques.^7^

Transitions from reversible to irreversible neuronal injury occur on timescales ranging from minutes to hours and even days. In vitro experiments have shown that the severity and reversibility of hypoxia-induced neuronal network damage depends on both the duration and depth of hypoxia.^16–19^ Based on these studies, it has been suggested that there are two separate pathophysiological pathways that contribute to brain injury: (1) functional network damage, resulting from persistent synaptic failure due to long-lasting but relatively mild hypoxia,^6,8^ and (2) structural tissue damage, resulting from neuronal membrane failure, swelling, and destruction due to relatively severe hypoxia.^13^

Combinations of functional and structural brain damage ultimately lead to persistent coma or clinical recovery, but it remains unclear how each of these mechanisms contribute to outcome, and whether both mechanisms can be independently assessed in vivo. So far, functional brain damage has been quantified using clinically scored EEG patterns (e.g., suppression or synchronous activity), quantitative EEG measures (e.g., alpha-to-delta ratio, discontinuity, or delta-band coherence), and functional connectivity within or between resting-state networks measured with resting-state functional MRI (rsfMRI).^20–22^ Structural tissue injury has been studied using structural MRI, e.g., by analysing changes seen on diffusion weighted imaging (DWI) or fluid attenuated inversion recovery (FLAIR) imaging, likely reflecting cytotoxic or vasogenic oedema. These studies have shown that patients with poor outcomes have more oedema, both visually^23,24^ and quantitatively as measured by the amount of diffusion restriction.^23–26^ However, the complex, underlying relationships between modalities have not yet been investigated, making it difficult to know which mechanisms are the drivers of post-anoxic encephalopathy after cardiac arrest, and how each of these mechanisms contributes to clinical neurological recovery.

Here, we address this issue by using a modular Bayesian framework (linked independent component analysis (LICA) model) designed to simultaneously model and discover common features across both structural (T1-weighted, DTI) and functional (rsfMRI) MRI modalities, to test which multimodal components relate to persistent coma or clinical recovery in a large sample (N=80) of patients with coma after cardiac arrest. Based on in vitro studies, we hypothesize that independent functional and structural mechanisms contribute to neurological outcome.

## Methods

### Study design

We performed a multicentre longitudinal cohort study in three Dutch hospitals: Rijnstate (Arnhem), Radboud university medical centre (Radboudumc, Nijmegen), and Maastricht University Medical Centre (MUMC+, Maastricht). Inclusion took place from June 2018 through March 2025. Data from patients included until November 2023 were used for current analyses.

### Study population

Patients were enrolled at the ICU within 72 hours of cardiac arrest. Inclusion criteria were: age ≥ 18 years, Glasgow Coma Scale (GCS) ≤ 8 on admission, and cardiac arrest due to cardiac cause or pulmonary embolism. Exclusion criteria were: pregnancy, life expectancy < 24 hours after admission, known progressive neurological disease (e.g. brain tumor or neurodegenerative disease), contraindication for MRI scanning, and pre-existing dependency in daily living.

### Treatment

Patients were monitored and treated according to local protocols and international guidelines for comatose patients after cardiac arrest.^27^ Targeted temperature management at 33-36 °C was initiated as soon as possible after arrival at the ICU according to local protocol and maintained for 24 hours. After 24 hours, passive rewarming was controlled and normothermia was actively maintained during at least 72 hours or until recovery of consciousness. Patients generally received a combination of propofol, midazolam, and morphine for sedation and analgesia.

Withdrawal of life-sustaining treatment (WLST) was considered at ≥ 72 hours after cardiac arrest, when patients were normothermic, and not receiving sedatives. Decisions on WLST were based on European guidelines, including bilateral absent somatosensory evoked potentials (SSEPs), treatment-resistant myoclonus, incomplete return of brainstem reflexes, and a highly malignant EEG. Brain MRI scans were never included in WLST decisions and treating physicians were blinded to the MRI measures published here.

### Outcome

The primary outcome measure was neurological outcome as defined by the cerebral performance category (CPC) score at six months after cardiac arrest. CPC scores were assessed by a standardized telephone interview based on the EuroQol-6D questionnaire. Neurological outcome was dichotomized as good (CPC 1-2: no to moderate neurological disability) or poor (CPC 3-5: severe neurological disability, vegetative state, or (brain) death).

### MRI data acquisition

Patients underwent MRI scanning on a 3T scanner, either Philips Ingenia (Rijnstate, MUMC+) or Siemens Skyra (Radboudumc). Scans were performed on day 2-9 after cardiac arrest. Scanning parameters are shown in **Supplementary Table S1**. For our current analyses we used T1-weighted, DWI, and rsfMRI scans.

### MRI data analyses

#### Feature Estimation

##### T1-weighted data

T1-weighted data were bias field corrected for intensity non-uniformity, rigid-body aligned, and segmented using CAT12.^28^ The resulting Jacobian deformation (JD) and grey matter volume (GMV) data were then smoothed with a 6 mm full-width half-maximum (FWHM)-kernel. Smoothed JD and GMV images were z-stratified and used for further analysis. Total intracranial volume was extracted from the CAT12 output to use as a covariate.

##### Diffusion-weighted Imaging data

DWI data were denoised and Gibbs ringing artifacts were removed using MRtrix3.^29^ Eddy current and bias field corrections were performed using FSL’s eddy tool^30^ and ANTs’ bias correction algorithm,^31^ respectively. We performed free water correction by modelling two separate tensors for freely diffusing extracellular water (fixed isotropic tensor; free water compartment) and the remaining signal (i.e. signal originating from restricted water; tissue compartment).^32^ Subsequently, free-water corrected FA and MD maps were calculated from the tissue compartment tensor. FA and MD maps were registered to standard space and FA maps were skeletonized using FSL’s Tract-Based Spatial Statistics Pipeline.^33^ Whole brain MD maps were used since previous studies showed importance of MD in grey matter in outcome prediction after cardiac arrest.^25^ MD maps were smoothed (kernel size σ = 2). Free water corrected whole brain MD and skeletonized FA maps were z-stratified and used for further analysis.

##### Resting-state functional MRI data

Resting-state fMRI scans were preprocessed using fMRIPrep 21.0.1.^34^ This includes motion correction, resampling to standard space, removal of non-steady state volumes, spatial smoothing with an isotropic, Gaussian kernel of 6 mm FWHM, and non-aggressive denoising based on independent component analysis (ICA-AROMA). Subsequently, we performed additional smoothing to reach a net 8 mm FWHM, white matter and cerebral spinal fluid nuisance regression, removal of first five volumes, and high pass filtering (0.007 Hz).

To find resting-state network (RSN) maps on a group-level, we first performed probabilistic ICA on a group level using FSL’s MELODIC.^35^ This resulted in twenty group-average spatially independent components. Each component was visually classified as RSN or noise. The RSN components were regressed to the individual maps using FSL’s Dual Regression,^36^ resulting in patient-specific connectivity maps for each RSN. Z-stratified connectivity maps of the visual network (VN), salience network (SN), frontoparietal network (FPN), executive control network (ECN), default-mode network (DMN), cerebellar network (CBN), somatosensory/motor network (SMN), and auditory network (AN) were used for further analysis.

#### Multimodal integration

##### Linked independent component analysis

To gain deeper insight into cross-modality patterns in a data-driven manner, we used LICA.^37^ This approach allows the integration of T1-weighted imaging, diffusion tensor imaging (DTI), and rsfMRI by extracting components that shared variance across these modalities. These independent components are statistically independent patterns of variation in the data, each representing a distinct spatial map that highlights brain regions where measures from different modalities co-vary across subjects. LICA achieves this by using subject-specific spatial maps derived from modality-specific input parameters. By enabling the combined processing of these modalities, LICA can identify common patterns of variation, revealing underlying relationships that may be missed when modalities are analysed in isolation.^38^ As a data-driven technique, LICA has the potential to reveal novel interactions and broader patterns that may not emerge in traditional hypothesis-driven studies. An overview of the complete analysis pipeline can be found in **Figure 1**.

**Figure 1.**
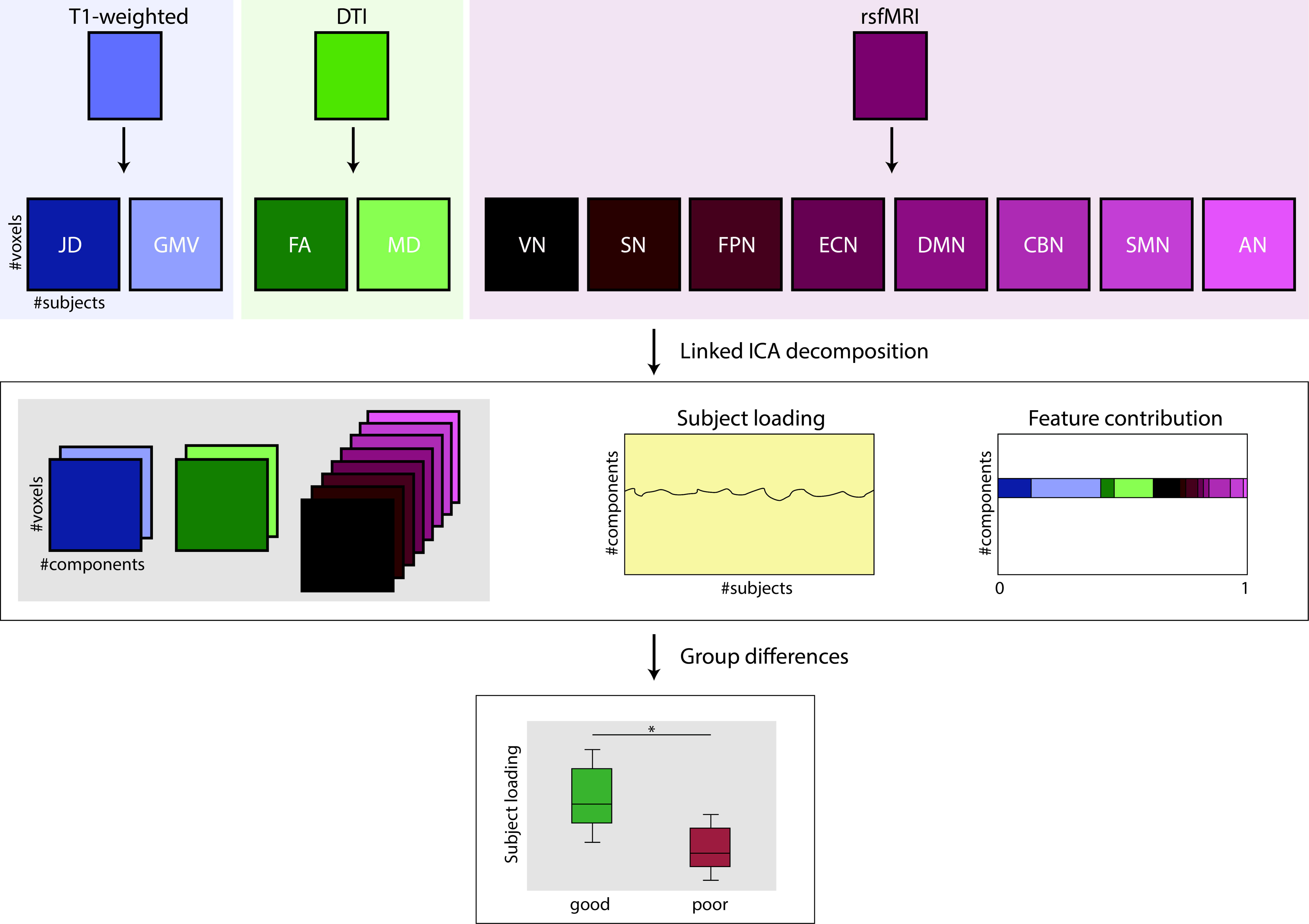
Overview of analysis pipeline. The individual modalities were preprocessed and spatial maps were extracted. A linked ICA decomposition was performed, resulting in 20 independent components. Subject loadings on the components were compared between patients with good and poor neurological outcomes. Adapted from Llera et al.^46^. AN = auditory network, CBN = cerebellar network, DMN = default-mode network, DTI = diffusion tensor imaging, ECN = executive control network, FA = fractional anisotropy, FPN = frontoparietal network, GMV = grey matter volume, JD = Jacobian deformation, MD = mean diffusivity, rsfMRI = resting-state functional MRI, SMN = somatosensory/motor network, SN = salience network, VN = visual network.

We combined the preprocessed and z-stratified JD, GMV, FA, MD, VN, SN, FPN, ECN, DMN, CBN, SMN, and AN maps in the LICA model. We ran the analysis with 20 components (sample size/4) and 1000 iterations.^37,38^ The subject loadings of the resulting components were used for statistical analyses. Spatial maps are thresholded at 2<|*z*|<5. Modalities contributing at least 5% to the independent components were considered for further evaluation.

### Statistical analyses

Data are presented as mean ± standard deviation (SD) or median [interquartile range (IQR)]. Group differences were assessed using chi-squared tests for ordinal variables and unpaired t-tests or Mann-Whitney U tests for continuous variables. Effect sizes were calculated using Cohen’s d. Correlation analyses were performed using Pearson’s r.

Subject loadings were corrected for age and sex by performing linear regression. The residuals after regression were used for statistical analyses. Group differences between good and poor neurological outcome were False-Discovery Rate (FDR) corrected for the number of components tested (*n* = 20). Components with significant differences between patients with good and poor outcomes or effect sizes ≥ 0.5 were further evaluated.

Relationships between components are presented in a descriptive way.

P-values < 0.05 after FDR-correction were assumed statistically significant. Statistical analyses were performed using R version 4.0.0.

### Standard protocol approvals, registrations, and patient consents

The medical ethics committee Arnhem-Nijmegen approved the study protocol, and the study protocol is registered (Cracking Coma study, ClinicalTrials.gov identifier: NCT03308305). Written informed consent was primarily obtained from a legal representative and, in case of recovery of capacity, later confirmed by the patient according to the Declaration of Helsinki. The study protocol and statistical analysis plan are available in eSAP 1.

### Data availability

Data that support the findings of this study are available from the corresponding author upon reasonable request.

## Results

We used data of eighty patients. Demographics and cardiac arrest related parameters are shown in **Table 1**. Patients with poor outcomes were older, had a non-shockable first heart rhythm more frequently, had longer times to ROSC, were more often sedated during the MRI scan, and had an absent N20 response more often.

**Table 1.** Demographics and cardiac arrest parameters of included patients.

|  | **Good outcome**<br>**(n = 49)** | **Poor outcome**<br>**(n = 31)** | **p-value** |
| --- | --- | --- | --- |
| **Male** | 41 (84%) | 22 (71%) | 0.283 |
| **Age (years)** | 59 ± 12 | 66 ± 11 | **0.010\*** |
| **OHCA** | 49 (100%) | 31 (100%) | 1.000 |
| **Shockable first heart rhythm** | 49 (100%) | 22 (71%) | **<0.001\*** |
| **Time to ROSC (minutes)** | 13 (IQR: 10-15) | 20 (IQR: 14-26) | **<0.001\*** |
| **Sedated during MRI scan** | 16 (33%) | 25 (81%) | **<0.001\*** |
| **Time from cardiac arrest to MRI (hours)** | 76 (IQR: 53-99) | 78 (IQR: 67-92) | 0.980 |
| **Absent SSEP N20 response** | 0 (0%) | 5 (16%) | **<0.001\*** |
| **EEG categories** |  |  | **<0.001\*** |
| **Continuous < 12 hours** | 19 (39%) | 1 (3%) |  |
| **Suppressed with/without superimposed synchronous activity > 24 hours** | 0 (0%) | 3 (10%) |  |
| **Other EEG pattern** | 30 (61%) | 27 (87%) |  |

Data are presented as number (%), mean ± standard deviation, or median (interquartile range). P-values < 0.05 were assumed statistically significant.

### Independent components

The twenty independent components are shown in **Figure 2**. Each component is composed of different combinations of data from structural and functional sequences. The relative contributions of the different structural and functional sequences is presented by different colours for each component.

**Figure 2.**
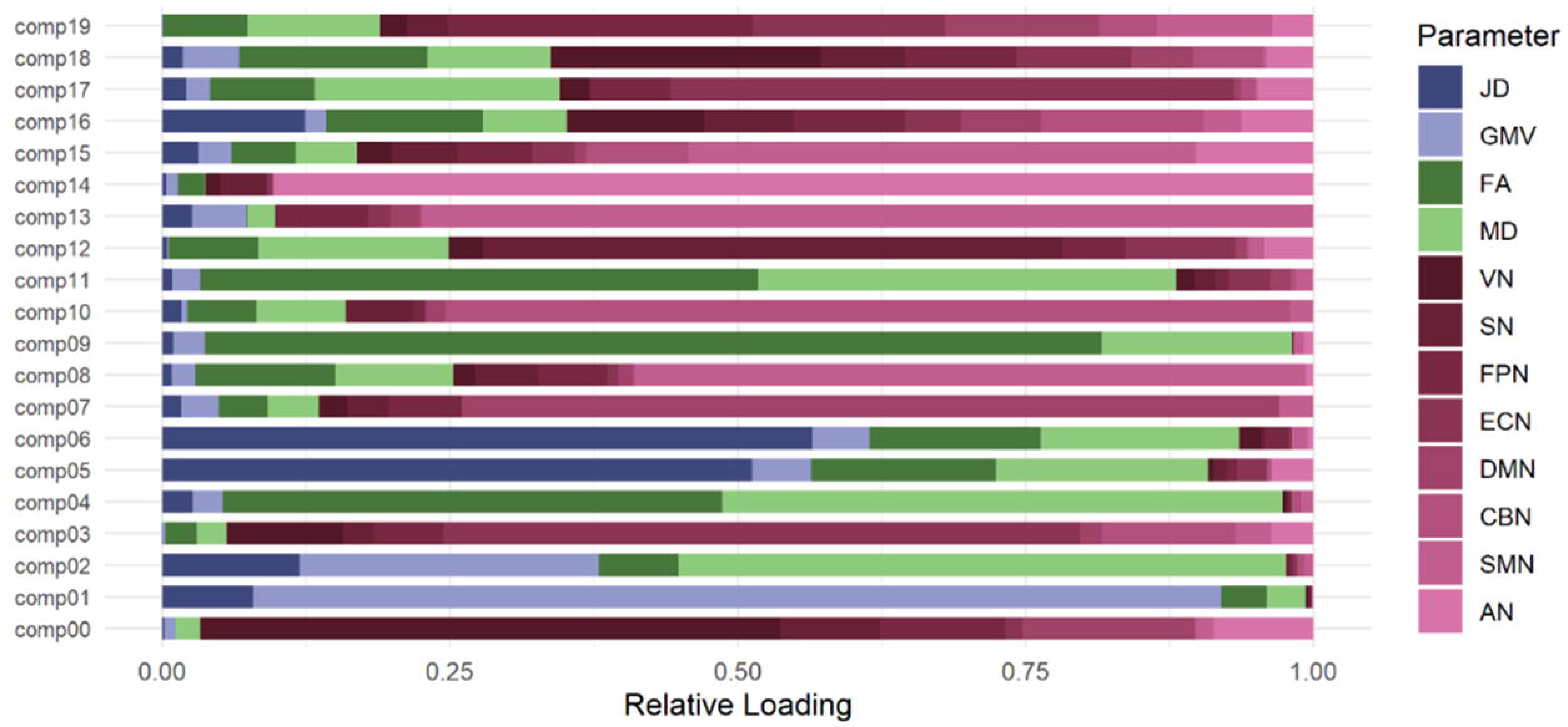
Relative loadings of parameters to components. AN = auditory network, CBN = cerebellar network, DMN = default-mode network, ECN = executive control network, FA = fractional anisotropy, FPN = frontoparietal network, GMV = grey matter volume, JD = Jacobian deformation, MD = mean diffusivity, SMN = somatosensory/motor network, SN = salience network, VN = visual network.

From the twenty components, six contained mainly functional information (components 00, 03, 07, 10, 14, and 15), seven contained mainly structural information (components 01, 02, 04, 05, 06, 09, and 11), five contained information from both functional and structural modalities (components 12, 16, 17, 18, and 19), and two were mainly dominated by information of one patient (> 50%, components 08 and 13). These two were not taken into account in subsequent analyses. Four components showed significant differences and/or medium to large effect sizes in subject loadings between patients with good and poor outcomes (FDR-corrected *p* < 0.05 or effect size > 0.5) and were all dominated by either functional (components 00 and 07) or structural (components 01 and 02) information. None of the components that contained information from both function and structure differed between outcome groups.

### Structural components relating to outcome

Component 02 showed the largest effect size for the difference between patients with good and poor neurological outcome (FDR-corrected *p* = 0.022, effect size 1.03), with lower loadings in patients with poor outcomes. The relative contributions of the different modalities were 53% for MD, 26% for GMV, 12% for JD, 7% for FA, and less than 5% for the other modalities. The bottom panel of **Figure 3** shows the results for this component. MD contributes positively to this component, and positive loadings are present throughout the cortex and deep grey nuclei (including the caudate, putamen, pallidum, hippocampus, amygdala, thalamus, and insula). This indicates that patients with poor outcomes have lower MD values in these regions. Both T1-weighted modalities have a predominantly negative contribution to the component, indicating that patients with poor outcome have larger volumes of several cortical regions (including the precuneus and precentral gyrus) and deep grey nuclei (including the caudate, putamen, pallidum, amygdala, thalamus, and insula). In addition, the JD has positive loadings for the ventricles and sulci, indicating smaller volumes of these regions in patients with poor outcome. The FA map contributes negatively to the component, most clearly in voxels in the fornix and thalamic radiation. This indicates that patients with poor outcome have higher FA values in these regions compared to patients with good outcomes. Subject loadings of component 2 were not related to sedatives during the MRI (uncorrected *p* = 0.176) or study site (uncorrected *p* = 0.397).

**Figure 3.**
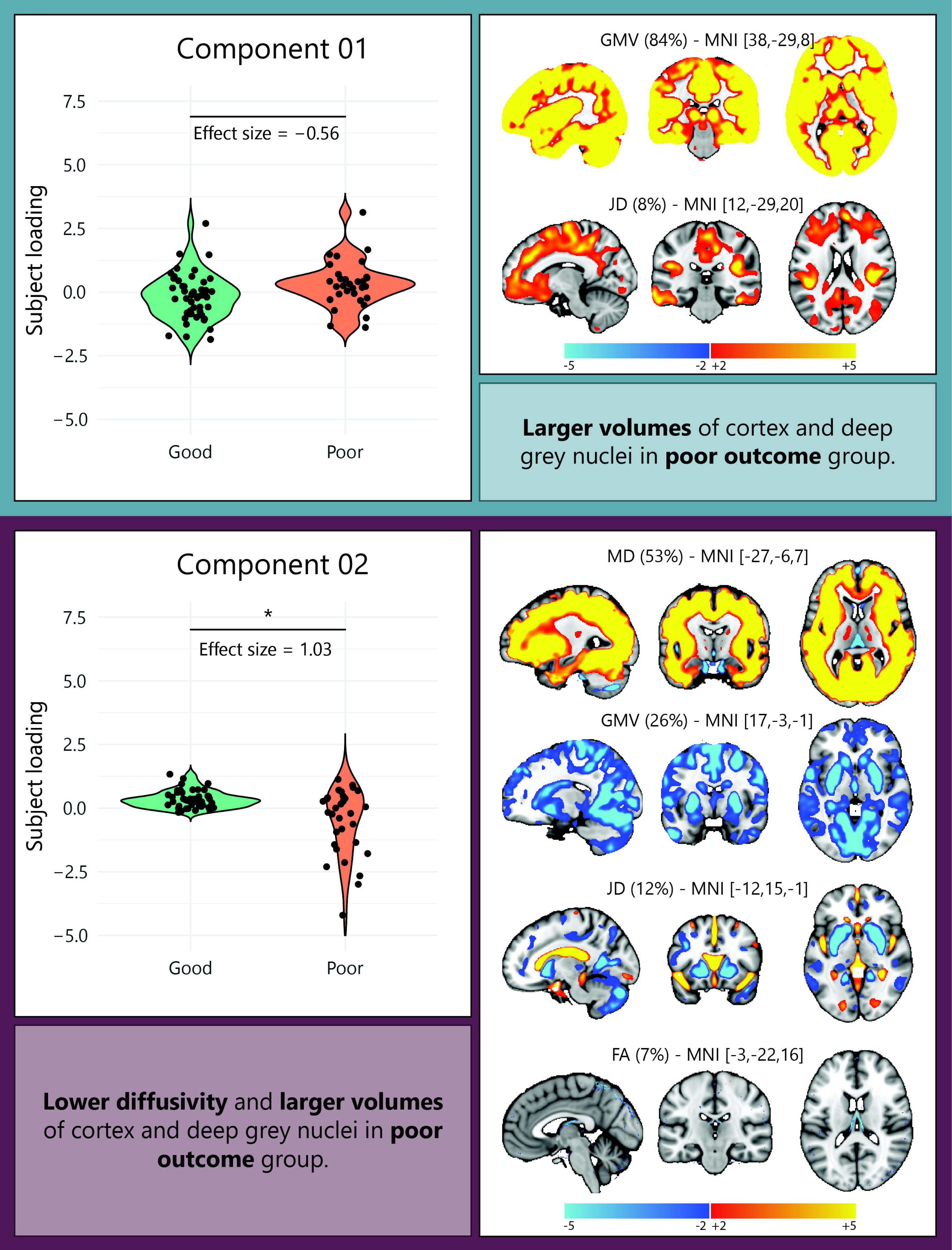
Summary of results of two components that include mainly structural data related to neurological outcome. Violin plots show the differences in subject loadings between patients with good and poor outcomes and spatial maps show the voxels within each modality that contributed to the specific component. FA = fractional anisotropy, GMV = grey matter volume, JD = Jacobian deformation, MD = mean diffusivity. Asterisks indicate significance levels (* *p* < 0.05).

Component 01 showed a borderline significant difference between patients with good and poor outcomes, but had a medium effect size (FDR-corrected *p* = 0.056, effect size -0.560) with higher loadings for patients with poor outcomes. The relative contributions of the modalities were 84% for GMV, 8% for JD, and less than 5% for the other modalities. The top panel of **Figure 3** shows the results for this component. GMV contributes positively to this component and significant loadings are present throughout the grey matter. Positive loadings of the JD are visible in the parietal operculum, cingulate gyrus, precuneus cortex, temporal fusiform cortex, and frontal pole. This indicates that patients with poor outcomes tend to have larger volumes of total cortex and deep grey nuclei compared to patients with good outcomes. Subject loadings of component 1 were not related to sedatives during the MRI (uncorrected *p* = 0.088) or study site (uncorrected *p* = 0.397).

### Functional components relating to outcome

Component 00 showed the largest difference between patients with good and poor neurological outcome (FDR-corrected *p* = 0.005, effect size 0.88), with lower loadings in patients with poor outcomes. The relative contributions of the different modalities were 50% for VN, 15% for DMN, 11% for FPN, 9% for SN, 9% for AN, and less than 5% for the other modalities. The top panel of **Figure 4** shows the results for this component. All five functional networks contribute positively to this component, and positive loadings are present throughout the networks. This indicates that patients with poor outcome have lower connectivity in the specific networks, without localization to specific parts of the networks. Subject loadings of component 0 were lower in patients who were sedated during the MRI compared to patients who weren’t sedated in both groups (uncorrected *p* < 0.001), but did not differ between study sites (uncorrected *p* = 0.335).

**Figure 4.**
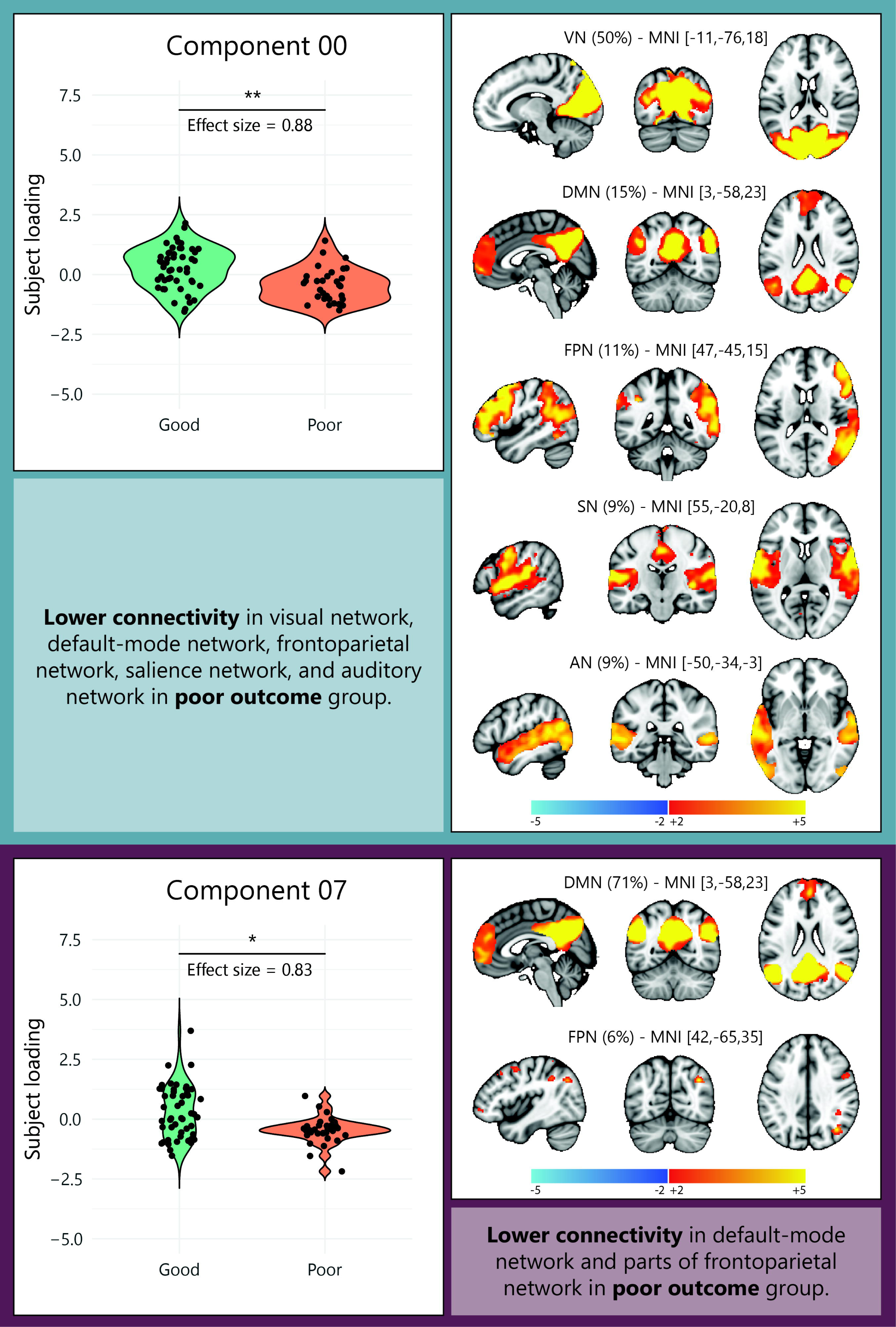
Summary of results of the two components that include mainly functional data related to neurological outcome. Violin plots show the differences in subject loadings between patients with good and poor outcomes and spatial maps show the voxels within each modality that contributed to the specific component. AN = auditory network, DMN = default-mode network, FPN = frontoparietal network, SN = salience network, VN = visual network. Asterisks indicate significance levels (* *p* < 0.05, ** *p* < 0.01).

Patients with poor outcome also showed lower subject loadings for component 07 (FDR-corrected *p* = 0.020, effect size 0.83). The relative contributions of the different modalities were 71% for DMN, 6% for FPN, and less than 5% for the other modalities. The bottom panel of **Figure 4** shows the results for this component. Both networks contribute positively to the component. The DMN shows positive contributions from voxels throughout the network, while the positive contributions from voxels within the FPN are localized in the dorsolateral prefrontal cortex, inferior frontal gyrus, inferior parietal gyrus, and premotor cortex. This indicates that patients with poor outcomes show reduced connectivity in the DMN and parts of the FPN. Subject loadings of component 7 were also lower in patients who were sedated during the MRI (uncorrected *p* = 0.002), but did not differ between study sites (uncorrected *p* = 0.318).

### Relationship between function and structure

We examined the relationship between the functional (component 07) and structural (component 02) components that showed the largest spread in data. **Figure 5** shows this relationship. Eyeballing of this plot reveals four quadrants, seemingly corresponding with the amount of brain damage. Relatively normal function *and* structure was observed in 22 patients with good outcome (45%) and 2 patients with poor outcome (6%), while functional impairment without structural injury was seen in 27 patients with good outcome (55%) and 16 patients with poor outcome (52%), and both functional and structural injury was seen in 0 patients with good outcome and 13 patients with poor outcome (42%). Notably, we did not observe any cases of structural injury without functional impairment in any outcome group. We used filled markers for patients with a reliable outcome prediction based on EEG (continuous<12h for good outcome and suppressed/synchronous>24h for poor outcome) and/or SSEP (absent N20 for poor outcome), while non-filled markers represent patients without a reliable prediction from these markers.

**Figure 5.**
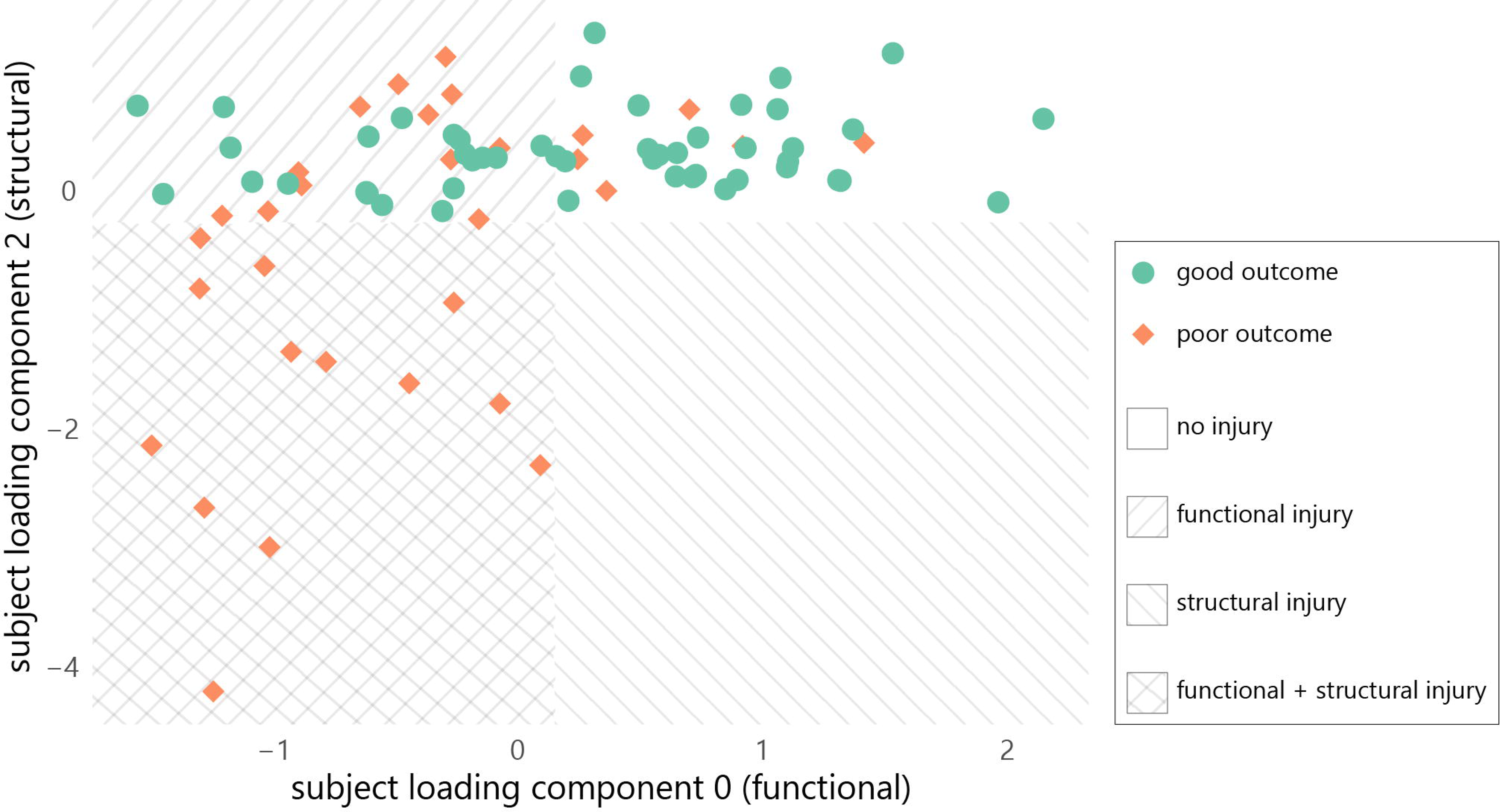
Mutual relationship between strongest functional (component 0) and structural (component 2) components and neurological outcome. Red diamonds correspond to patients with poor outcome, green circles to patients with good outcome. Plot is divided into four quarters: no structural and no functional injury (right upper quadrant), functional injury without structural injury (left upper quadrant), functional and structural injury (right lower quadrant), and structural injury without functional injury (right lower quadrant). Thresholds for the four quadrants are based on eyeballing of the scatterplots.

## Discussion

We parsed aspects of the pathophysiological mechanisms contributing to persistent coma or clinical recovery of patients with postanoxic encephalopathy after cardiac arrest using a linked independent component analysis approach to model and discover common features across multiple structural and functional MRI modalities. We found two independent functional and two structural determinants of good and poor outcome. Structural changes contributing to poor neurological outcome consisted of lower MD and larger volumes of the cortex and deep grey nuclei, likely reflecting a combination of cytotoxic and vasogenic oedema.^39,40^ Functional changes related to poor neurological outcome consisted of reduced functional connectivity in several resting-state networks, most prominently the VN, DMN, and FPN, which probably reflects decreased synaptic connectivity.^41^ None of the components relating to outcome captured a relationship between structural tissue and functional network changes, pointing towards divergent pathways toward persistent brain damage or recovery after cardiac arrest.

Functional network impairment was observed in both outcome groups, with approximately 50% of patients with either good or poor outcome showing functional network impairment *without* signs of structural tissue damage. This is consistent with findings from studies under experimental in vitro and in vivo conditions, showing that impairments in functional neuronal network activity can occur in the absence of membrane failure or cell swelling during and after transient mild to moderate hypoxia.^6,8,17^ Clinically, this mirrors observations in post-cardiac arrest patients, where pathological EEG patterns (e.g., suppressed, low voltage, or burst suppression) often normalize on time scales of hours during ICU admission.^11,20^ These patterns most likely reflect synaptic failure,^6,19^ which may be reversible if blood flow is restored in time.^6,10^ However, prolonged hypoxia may result in persistent synaptic dysfunction^16,17^, even with intact membrane potential and without cell swelling. Most experimental evidence supports that early synaptic failure is located pre-synaptically and caused by impaired neurotransmitter release.^6,9,10^ Mechanisms underlying the failure of transmitter release include dysfunction of presynaptic ATP-dependent calcium channels^42^ and impaired docking of glutamate containing vesicles due to impaired phosphorylation.^8^ This is a likely explanation of our observed functional network changes without structural tissue damage, especially in patients with a poor outcome.

Approximately 40% of our poor outcome patients and none of the good outcome patients had signs of structural brain tissue changes, including decreased diffusivity and increased volume of the cerebral cortex and deep grey nuclei, probably reflecting oedema. In relatively severe hypoxia, synaptic impairment may be accompanied by membrane failure, i.e., loss of ion gradients across the plasma membrane. As a result, intracellular osmolality increases, and water enters the cells. This leads to the development of cytotoxic oedema^13,14^ which is captured by lower MD. As the ischemic period progresses, the integrity of the blood-brain barrier is compromised, allowing osmotically active substances and water to accumulate in the extracellular space. This leads to vasogenic oedema^13^, which is seen as increased volume on T1-weighted imaging. Indeed, our structural components showed a combination of diffusion restriction (sign of cytotoxic oedema^39^) and volume increase (sign of vasogenic oedema^40^). Structural tissue injury was observed exclusively in patients with poor outcomes, reinforcing its association with severe hypoxia. However, unlike our finding, previous studies have shown that structural tissue damage may also be present in patients with good outcomes, mostly showing as mild changes on DWI or FLAIR scans.^23,43^ We expect these lesions in good outcome patients to be either partially reversible or located such that they do not significantly affect overall brain function.

As expected, most patients with no evidence of functional or structural impairment had a good neurological outcome. However, we found two patients with poor outcome in this group. Both had a CPC score of 3, indicating recovery of consciousness with severe disability. CPC 3 indicates a certain amount of postanoxic encephalopathy, which we apparently did not detect by our MRI measures.

Our individual findings on associations between functional, diffusion, and volumetric measures and neurological outcome are mostly consistent with previous research, showing low connectivity in resting-state networks^22,44^ and low diffusivity in patients with a poor outcome.^23,25,26,43^ The results for volumetric measures differ from a previous study showing that patients with poor outcome had thinner frontal cortices and smaller volumes of the thalamus, putamen, and pallidum,^45^ whereas we found that larger cortical volume and larger deep grey matter nuclei volumes were associated with poor outcome. These differences may be explained by the timing of the MRI scans. Our MRI scans about 3 days after cardiac arrest probably captured oedema, whereas MRI scans collected in previous work at an average of 16 days likely showed the secondary effects of atrophy after brain injury. The higher FA values in patients with poor outcome, particularly in regions near the ventricles, may be explained by residual effects of free water compartments and the presence of enlarged ventricles, despite the applied free water correction.

Several components not associated with neurological outcome contained information from both functional and structural modalities. This finding suggests a link between brain function and structure. However, since combined components lacked a relationship with good or poor outcome, these most likely represent effects unrelated to brain damage resulting from the cardiac arrest. We assume that these components may reflect baseline inter-individual differences, rather than injury. Factors such as age, sex, or other demographic or physiological inter-individual factors are known to influence both structural and functional brain characteristics and probably contribute to these components.^46,47^

Our findings have potential implications for the management of postanoxic encephalopathy. Although we demonstrate different pathomechanisms leading to persistent coma or recovery, currently all patients are lumped together and treated the same. It is unlikely that isolated, reversible synaptic failure warrants the same neuroprotective strategy as cell and tissue swelling with excitotoxicity and blood brain barrier disruption. For example, the effects of mild therapeutic hypothermia were different in patients with EEG patterns indicating different degrees of encephalopathy.^48^ Over the past decades, more than twenty potential neuroprotective treatments have been tested in clinical trials, but none has shown unequivocal evidence of efficacy.^49^ An important rationale behind all studied strategies, including the widely used hypothermia, was the prevention of secondary tissue damage, either by inhibiting neuronal activity to conserve remaining energy to maintain basic cellular processes, or by inhibiting potentially detrimental steps in the pathophysiological cascade. This implies that previously tested treatment modalities have mainly targeted processes associated with ongoing tissue changes, such as those present in patients with a relatively poor prognosis. Isolated synaptic failure probably warrants a different approach. For example, non-invasive brain stimulation techniques have been associated with increased recovery of physiological neuronal activity^18^ and improved synapse recovery in experimental models of hypoxia induced synaptic failure.^18,50^

Strengths of this study include the prospective study design, the data-driven analysis approach without a priori hypotheses about specific brain regions or networks, and the use of a linked ICA model that is more sensitive to complex combinations of functional and structural brain patterns than unimodal analyses. To our knowledge, we are the first to study interdependencies between T1-weighted, DTI, and rsfMRI in this patient population.

Our study also has several limitations. First, sedation may have influenced our results. Our functional components were significantly related to sedation. However, we decided to not correct for this effect, because sedation is also associated with neurological outcome. Patients with good outcomes typically require either high levels of sedation or no sedation at all, because of recovered consciousness, whereas patients with poor outcomes generally receive moderate sedation during the scan. Second, we cannot completely rule out the effects of self-fulfilling prophecy. However, decisions to withdraw life-sustaining treatment were never based on brain MRI, and treating physicians did not have access to the quantitative measures we used for this analysis. Third, we used data from different study sites and different MRI vendors. This may have affected our results. Nonetheless, the subject loadings of the four relevant components identified in this paper did not differ between study sites, suggesting that the influence of this covariate in our analysis was limited. Fourth, we did not include a healthy control group. Because we were interested in differences between patients with good and poor outcomes, we did not compare our results with healthy controls. It might be interesting to do so and learn the differences between patients with postanoxic encephalopathy and healthy controls, but this was beyond the scope of out study. Finally, we did not include EEG data in our analysis. As a more direct measure of synaptic function than fMRI, EEG could provide valuable insights and should be considered in future research.

We conclude that both functional network and structural tissue injury are associated with poor neurological outcome after cardiac arrest, but they appear to represent two distinct cerebral mechanisms with different characteristics in terms of reversibility. Future research may focus on unravelling the specific mechanisms underlying these differences, with an emphasis on identifying interventions that can minimize persistent network failure and enhance functional recovery.

## Acknowledgements

The authors thank the staff of the ICU and radiology departments of the participating centres for constructive assistance in obtaining informed consent and performing MRI scans.

## Funding

Jeannette Hofmeijer: Dutch Heart Foundation (2018T070)

Rick C. Helmich: NOW (0915017201004)

Cracking coma and this analysis: institutional grants from University of Twente, Radboudumc, and Rijnstate hospital

## Competing interests

The authors report no competing interests.

## Notes

### Competing Interest Statement

The authors have declared no competing interest.

### Author Declarations

Medical ethics committee Arnhem-Nijmegen gave ethical approval for this work

