## Supplementary information for "Separate functional and structural cerebral mechanisms relate to postanoxic coma recovery"

**Table S1**. Overview of scanning parameters for the T1-weighted, DTI, and rsfMRI scans on both the Philips Ingenia and Siemens Skyra scanners.

| | T1-weighted | | DTI | | rsfMRI | |
|---|---|---|---|---|---|---|
| | Philips | Siemens | Philips | Siemens | Philips | Siemens |
| TE (ms) | 3.8 | 3.41 | 9500 | 9700 | 27 | 27 |
| TR (ms) | 8300 | 2400 | 95 | 95 | 2220 | 2280 |
| Voxel size (mm) | 1.0*1.0*1.0 | 0.9*0.9*1.0 | 2.0*2.0*2.0 | 2.0*2.0*2.0 | 3.0*3.0*3.0 | 3.2*3.2*3.0 |
| B0-images (#) | | | 1 | 1 | | |
| B1000-images (#) | | | 32 | 30 | | |
| Volumes (#) | | | | | 220 | 220 |

*DTI = diffusion tensor imaging, rsfMRI = resting-state functional MRI, TE = echo time, TR = repetition time*
